# Machine learning algorithm to predict fragility fractures and identification of important features – an explainable approach

**DOI:** 10.1101/2025.07.10.25331257

**Authors:** Sayem Borhan, Alexandra Papaioannou, Jonathan Adachi, Shrey Acharya, Suzanne N. Morin, David Goltzman, David A. Hanley, Claudie Berger, Lehana Thabane, Parminder Raina

## Abstract

In this study, we developed ML algorithms to predict fragility fractures, considering the occurrence of fractures at different skeletal sites. We investigated seven ML algorithms (LASSO, Elastic Net, Random Forest, Decision Tree, Neural Network, XGBoost and Logistic Regression) using the data from the Canadian Multicentre Osteoporosis Study (CaMos) with participants aged 50 years or older. We considered 73 baseline features, including age, sex, menopause status, and bone mineral density (BMD), and the outcome was the first incidence of fracture at any of the following sites: hip, spine, pelvis, ribs, shoulder, and forearm, over a 19-year follow-up period. Data were divided into training (70%) and testing (30%) datasets. The ML algorithms were trained on the training dataset and evaluated on the test dataset in terms of the ROC_AUC. SHapley Additive exPlanations (SHAP) analysis was performed to identify the important features that contribute to the prediction of fracture, and to investigate the interaction among these features.

In total, 7,753 subjects were included in the study. Approximately 72% were female, and the average age was 67 years. We found that the XGBoost algorithm had a slightly better ROC_AUC (0.70; 95% CI: 0.67, 0.73). From the SHAP analysis, we found that BMD was the most important feature that contributed to the prediction. The other important features include age, previous fracture, osteoporosis and menopausal status. Total hip BMD interacted the most with femoral neck BMD, lumbar spine BMD interacted the most with weight, previous fracture status interacted the most with femoral neck BMD, and age interacted the most with lumbar spine BMD.

This study demonstrated that XGBoost was the most effective algorithm for predicting fragility fractures. In addition, we identified important features that contribute to the prediction of fragility fractures. Intervention focusing on these features will help to prevent the incidence of these fractures.

**Lay summaries:** We developed machine learning (ML) algorithms to predict fragility fractures, considering the incidence of fractures at different skeletal sites, including the hip, spine, pelvis, ribs, shoulder, or forearm, using 19 years of follow-up data from the Canadian Multicentre Osteoporosis Study (CaMos). We investigated seven ML algorithms and found that XGBoost had slightly better performance compared to other algorithms. We identified important factors that increase the risk of fractures, including BMD, age, and previous fracture. We also demonstrated how the interaction between these factors increases the risk of fractures. The intervention focusing on these factors will help to prevent fragility fractures.

## Introduction

Fragility fractures pose a significant health burden that predominately affects older adults, with a global burden of 178 million new documented cases, 25.8 million years lived with disability (YLD), and 455 million prevalent cases with acute or long-term symptoms^1^. These fractures occur from a “low trauma” that would otherwise not typically injure bone^2^ have significant detrimental quality of life (QoL) implications, including impaired mobility and institutionalization, and can occur at different skeletal sites, including hip, spine and forearm^3,4^. The hip and spine fractures have the most significant consequences, including a high 1-year mortality rate and markedly reduced independence^3,5–8^. It is estimated that these fractures will cost the American healthcare system 25.3 billion dollars by 2025^9^. Due to the high prevalence and the growing aging population^10^, the economic and QoL burden of fragility fractures will continue to increase over time^11^. Therefore, urgent measures are needed to prevent and mitigate these fractures.

Identifying individuals at a higher risk of fragility fractures is imperative to enable the implementation of early preventative strategies^10–12^, including lifestyle and behavioural changes to reduce the incidence of fractures^12,13^. FRAX^14^ is one of the most widely used predictive tools, and uses a statistical method to estimate the 10-year probability of a fracture. FRAX considers several factors, including bone mineral density (BMD), age, sex, height, weight, and previous fractures^14^. However, FRAX tends to underestimate fracture risk^15,16^ and relies on traditional statistical methods that assume no interaction between risk factors, which is not always accurate^17^. Its performance also varies across countries and does not account for race, ethnicity, or social determinants of health^18^. It is essential to increase the accuracy of the prediction by considering factors not included in FRAX and utilizing machine learning (ML) algorithms as an alternative to the statistical methods^19^.

The ML algorithms have the potential to improve the accuracy of the fragility fractures prediction and discern high-risk groups^18–23^. For instance, ML performs better than traditional logistic regression in predicting hip fracture^24,25^ and outperforms FRAX in fracture prediction^26,27^. The investigations of ML algorithms to predict fragility fractures varied in terms of data sources, type of ML algorithm, and type and number of features considered for the analysis^19,20^. Smets et al., in their systematic review, found that the majority of the studies (12 out of 14) focused on predicting only hip fracture^20^ and fracture was measured with a follow-up of less than 10 years^20^. Fractures can occur at different skeletal sites, including the hip, spine, forearm, pelvis and ribs, but evidence considering all these sites has been lacking. Vries et al. considered several major osteoporotic fractures, including hip, spine, wrist and humerus, in their study of ML algorithms^23^. However, this study was limited to participants with osteoporosis and osteopenia who had sustained a previous fracture.

In addition, we have limited evidence regarding the adoption of the explainable ML approach, which involves assessing and quantifying the contribution of features to the prediction of the outcome of interest^28^. It is also helpful to understand how the features interact with one another^29^ as it enables researchers to understand the condition and potential management approaches. The Shapely Additive Explanation (SHAP) analysis helps us to identify the importance of features and how these features interact with one another^28,30^. SHAP analysis is based on game theory that yields SHAP values to quantify the contribution of a feature and feature-feature interaction to the prediction of an outcome ^28,30^.

In this study, we adopted an explainable approach and investigated various ML algorithms to predict the incidence of fragility fractures considering multiple skeletal sites, which were defined as the first incidence of a fracture at any of hip, spine, pelvis, ribs, forearm, or shoulder over 19-years of follow-up data from the Canadian Multicentre Osteoporosis Study (CaMos)^31^. The primary objective of this study was to identify the best ML algorithm to predict the incidence of fragility fractures. The secondary objectives were to determine the important features that contribute to this prediction and how the important features interact with other features.

## Materials and Methods

### Setting and Study Population

This study was based on the Canadian Multicentre Osteoporosis Study (CaMos), a population-based longitudinal study where data were collected from over 9000 adults aged 25 and older, irrespective of osteoporosis status. Details of the CaMos can be found elsewhere^31^. Baseline assessments were conducted between 1995 and 1997, while the 19-year follow-up assessment was conducted in 2019^31^. In-person interviews were conducted at years 5, 10, and 16, while yearly data were collected through self-reported questionnaires^31^. We included participants aged 50 years or older (n=7,753).

### Fragility Fractures and Features

Fragility fractures were defined using the World Health Organization (WHO) definition, which classified them as “fractures caused by injury that would be insufficient to fracture normal bone” ^2^. We considered the first occurrence of any of hip, spine, pelvis, forearm, shoulder, or ribs fracture that occurred due to “low trauma,” such as falling from a standing height or less, over the 19 years of follow-up. Fractures were identified through interviews or questionnaires and subsequently confirmed through X-rays and/or medical reports^31^. Seventy-three baseline features were considered in this study, including age, sex, vitamin D and calcium intake, and bone mineral density (total hip, lumbar spine and femoral neck). The details of the features are shown in Table 1.

**Table 1:** Descriptive summary of the included features by fracture status.

| Features | Overall<br>(n=7,753) | No Fracture<br>(n=6,835) | Fracture*<br>(n=918) |
| --- | --- | --- | --- |
| Age, Mean (SD), y | 66.67 (9.35) | 66.38 (9.41) | 68.85 (8.56) |
| Sex, No. (%) |  |  |  |
| Female | 5566 (71.79) | 4785 (70.01) | 781 (85.08) |
| Male | 2187 (28.21) | 2050 (29.99) | 137 (14.92) |
| Education, No. (%) |  |  |  |
| <post-secondary | 5853 (75.50) | 5175 (75.72) | 678 (73.86) |
| ≥post-secondary | 1899 (24.50) | 1659 (24.28) | 240 (26.14) |
| Living alone, No. (%) |  |  |  |
| Yes | 2573 (33.19) | 2211 (32.35) | 362 (39.43) |
| No | 5180 (66.81) | 4624 (67.65) | 556 (60.57) |
| Living with another adult, No. (%) |  |  |  |
| Yes | 5119 (99.32) | 4569 (99.28) | 550 (99.64) |
| No | 35 (0.68) | 33 (0.72) | 2 (0.36) |
| Employed, No. (%) |  |  |  |
| Yes | 3146 (40.58) | 2800 (40.97) | 346 (37.6) |
| No | 4607 (59.42) | 4035 (59.03) | 572 (62.31) |
| Have regular doctor, No. (%) |  |  |  |
| Yes | 7516 (96.94) | 6618 (96.83) | 898 (97.82) |
| No | 237 (3.06) | 217 (3.17) | 20 (2.18) |
| Rheumatoid arthritis, No. (%) |  |  |  |
| Yes | 490 (6.51) | 409 (6.16) | 81 (9.09) |
| No | 7042 (93.49) | 6232 (93.84%) | 810 (90.91) |
| Osteoporosis, No. (%) |  |  |  |
| Yes | 703 (9.25) | 527 (7.86) | 176 (19.69) |
| No | 6900 (90.75) | 6182 (92.14) | 718 (80.31) |
| Osteoarthritis, No. (%) |  |  |  |
| Yes | 2529 (34.06) | 2217 (33.83) | 312 (35.78) |
| No | 4897 (65.94) | 4337 (66.17) | 560 (64.22) |
| Thyroid, No. (%) |  |  |  |
| Yes | 1157 (14.93) | 978 (14.32) | 179 (19.5) |
| No | 6590 (85.07) | 5852 (85.68) | 738 (80.48) |
| Liver disease, No. (%) |  |  |  |
| Yes | 192 (2.48) | 162 (2.38) | 30 (3.28) |
| No | 7542 (97.52) | 6656 (97.62) | 886 (96.72) |
| Scoliosis, No. (%) |  |  |  |
| Yes | 381 (4.96) | 335 (4.94) | 46 (5.07) |
| No | 7306 (95.04) | 6445 (95.06) | 861 (94.93) |
| Eating disorder, No. (%) |  |  |  |
| Yes | 28 (0.36) | 21 (0.31) | 7 (0.76) |
| No | 7722 (99.64) | 6811 (99.69) | 911 (99.24) |
| Breast cancer, No. (%) |  |  |  |
| Yes | 264 (3.41) | 223 (3.27) | 41 (4.47) |
| No | 7,484 (96.59) | 6,607 (96.73) | 877 (95.53) |
| Uterine cancer, No. (%) |  |  |  |
| Yes | 135 (1.74) | 109 (1.60) | 26 (2.84) |
| No | 7608 (98.26) | 6718 (98.40) | 890 (97.16) |
| Prostate cancer, No. (%) |  |  |  |
| Yes | 89 (1.15) | 80 (1.17) | 9 (0.98) |
| No | 7,659 (98.85) | 6,750 (98.83) | 909 (99.02) |
| Kidney disease, No. (%) |  |  |  |
| Yes | 733 (9.46) | 643 (9.41) | 90 (9.80) |
| No | 7018 (90.54) | 6190 (90.59) | 828 (90.20) |
| IBD, No. (%) |  |  |  |
| Yes | 436 (5.65) | 362 (5.32) | 74 (8.12) |
| No | 7,276 (94.35) | 6,439 (94.68) | 837 (91.88) |
| Heart disease, No. (%) |  |  |  |
| Yes | 577 (7.47) | 508 (7.47) | 69 (7.53) |
| No | 7144 (92.53) | 6297 (92.53) | 847 (92.47) |
| CVT/TIA, No. (%) |  |  |  |
| Yes | 354 (4.58) | 309 (4.53) | 45 (4.91) |
| No | 7376 (95.42) | 6505 (95.47) | 871 (95.09) |
| COPD, No. (%) |  |  |  |
| Yes | 684 (10.10) | 598 (10.08) | 86 (10.26) |
| No | 6085 (89.90) | 5333 (89.92) | 752 (89.74) |
| Diabetes, No. (%) |  |  |  |
| Yes | 606 (7.82) | 541 (7.92) | 65 (7.09) |
| No | 7146 (92.18) | 6294 (92.08) | 852 (92.91) |
| Neuromuscular, No. (%) |  |  |  |
| Yes | 229 (2.95) | 199 (2.91) | 30 (3.27) |
| No | 7524 (97.05) | 6636 (97.09) | 888 (96.73) |
| Phlebitis, No. (%) |  |  |  |
| Yes | 493 (6.38) | 417 (6.12) | 76 (8.29) |
| No | 7240 (93.62) | 6399 (93.88) | 841 (91.71) |
| PAGET, No. (%) |  |  |  |
| Yes | 16 (0.21) | 13 (0.19) | 3 (0.33) |
| No | 7724 (99.79) | 6812 (99.81) | 912 (99.67) |
| Gall bladder surgery, No. (%) |  |  |  |
| Yes | 1428 (18.42) | 1224 (17.91) | 204 (22.22) |
| No | 6325 (81.58) | 5611 (82.09) | 714 (77.78) |
| Intestine surgery, No. (%) |  |  |  |
| Yes | 404 (5.21) | 337 (4.93) | 67 (7.30) |
| No | 7349 (94.79) | 6498 (95.07) | 851 (92.70) |
| Parathyroid surgery, No. (%) |  |  |  |
| Yes | 20 (0.26) | 18 (0.26) | 2 (0.22) |
| No | 7733 (99.74) | 6817 (99.74) | 916 (99.78) |
| Stomach surgery, No. (%) |  |  |  |
| Yes | 254 (3.28) | 207 (3.03) | 47 (5.12) |
| No | 7499 (96.72) | 6628 (96.97) | 871 (94.88) |
| Thyroid surgery, No. (%) |  |  |  |
| Yes | 208 (2.68) | 179 (2.62) | 29 (3.16) |
| No | 7545 (97.32) | 6656 (97.38) | 889 (96.84) |
| Falls last month, No. (%) |  |  |  |
| Yes | 503 (6.49) | 424 (6.20) | 79 (8.61) |
| No | 7250 (93.51) | 6411 (93.80) | 839 (91.39) |
| Hip fracture at baseline, No. (%) |  |  |  |
| Yes | 144 (1.86) | 109 (1.60) | 35 (3.81) |
| No | 7605 (98.14) | 6722 (98.40) | 883 (96.19) |
| Pelvis fracture at baseline, No. (%) |  |  |  |
| Yes | 82 (1.06) | 60 (0.88) | 22 (2.40) |
| No | 7668 (98.94) | 6772 (99.12) | 896 (97.60) |
| Spine fracture at baseline, No. (%) |  |  |  |
| Yes | 178 (2.30) | 136 (1.99) | 42 (4.58) |
| No | 7571 (97.70) | 6696 (98.01) | 875 (95.42) |
| Forearm fracture at baseline, No. (%) |  |  |  |
| Yes | 1134 (14.64) | 937 (13.72) | 197 (21.51) |
| No | 6611 (85.36) | 5892 (86.28) | 719 (78.49) |
| Ribs fracture at baseline, No. (%) |  |  |  |
| Yes | 345 (4.45) | 283 (4.14) | 62 (6.75) |
| No | 7403 (95.55) | 6547 (95.86) | 856 (93.25) |
| Other fracture at baseline, No. (%) |  |  |  |
| Yes | 1430 (18.48) | 1202 (17.61) | 228 (24.92) |
| No | 6310 (81.52) | 5623 (82.39) | 687 (75.08) |
| Walk past 6 months, No. (%) |  |  |  |
| >5 hours | 1485 (19.16) | 1270 (18.58) | 215 (23.42) |
| ≤5 hours | 6267 (80.84) | 5564 (81.42) | 703 (76.58) |
| Work type, No. (%) |  |  |  |
| Not sitting | 5558 (71.80) | 4874 (71.42) | 684 (74.59) |
| Sitting | 2183 (28.20) | 1950 (28.58) | 233 (25.41) |
| Participate in physical activity, No. (%) |  |  |  |
| Yes | 4297 (55.42) | 3779 (55.29) | 518 (56.43) |
| No | 3456 (44.58) | 3056 (44.71) | 400 (43.57) |
| Strenuous sports, No. (%) |  |  |  |
| Yes | 966 (12.46) | 870 (12.73) | 96 (10.46) |
| No | 6785 (87.54) | 5963 (87.27) | 822 (89.54) |
| Vigorous work, No. (%) |  |  |  |
| Yes | 1370 (17.68) | 1251 (18.31) | 119 (12.96) |
| No | 6380 (82.32) | 5581 (81.69) | 799 (87.04) |
| Moderate activity, No. (%) |  |  |  |
| Yes | 1370 (17.68) | 1251 (18.31) | 119 (12.96) |
| No | 2895 (37.36) | 2581 (37.78) | 314 (34.20) |
| Sleep, No. (%) |  |  |  |
| ≥7 hours | 5463 (70.51) | 4860 (71.15) | 603 (65.76) |
| <7 hours | 2285 (29.49) | 1971 (28.85) | 314 (34.24) |
| Sun exposure, No. (%) |  |  |  |
| Yes | 2502 (32.28) | 2224 (32.55) | 278 (30.28) |
| No | 5249 (67.72) | 4609 (67.45) | 640 (69.72) |
| Race, No. (%) |  |  |  |
| White | 7405 (95.51) | 6509 (95.23) | 896 (97.60) |
| Non-white | 348 (4.49) | 326 (4.77) | 22 (2.40) |
| Calcium intake/day, Mean (SD), mg | 1020.60 (617.72) | 1013.78 (618.61) | 1071.56 (608.93) |
| Vitamin D intake/day, Mean (SD), mcg | 7.45 (25.09) | 7.09 (23.43) | 10.11 (34.92) |
| Menopause, No. (%) |  |  |  |
| Not menopausal | 2463 (31.77) | 2312 (33.83) | 151 (16.45) |
| Menopausal | 3393 (43.76) | 2907 (42.53) | 486 (52.94) |
| Both ovaries removed | 1897 (24.47) | 1616 (23.64) | 281 (30.61) |
| Ever smoker, No (%) |  |  |  |
| Yes | 4161 (53.68) | 3668 (53.68) | 493 (53.70) |
| No | 3590 (46.32) | 3165 (46.32) | 425 (46.30) |
| Drink, Mean (SD),y | 150.40 (304.89) | 151.78 (302.80) | 140.19 (320.01) |
| CVD history, No. (%) |  |  |  |
| Yes | 5494 (75.53) | 4837 (75.41) | 657 (76.40) |
| No | 1780 (24.47) | 1577 (24.59) | 203 (23.60) |
| Fracture history, No. (%) |  |  |  |
| Yes | 3721 (53.66) | 3248 (53.24) | 473 (56.78) |
| No | 3213 (46.34) | 2853 (46.76) | 360 (43.22) |
| Osteoarthritis history, No. (%) |  |  |  |
| Yes | 2737 (41.16) | 2407 (41.02) | 330 (42.25) |
| No | 3912 (58.84) | 3461 (58.98) | 451 (57.75) |
| Osteoporosis history, No. (%) |  |  |  |
| Yes | 916 (14.29) | 760 (13.41) | 156 (20.94) |
| No | 5495 (85.71) | 4906 (86.59) | 589 (79.06) |
| Scoliosis history, No. (%) |  |  |  |
| Yes | 337 (4.84) | 293 (4.78) | 44 (5.35) |
| No | 6621 (95.16) | 5843 (95.22) | 778 (94.65) |
| Breast cancer history, No. (%) |  |  |  |
| Yes | 979 (13.69) | 863 (13.70) | 116 (13.65) |
| No | 6172 (86.31) | 5438 (86.30) | 734 (86.35) |
| Uterine cancer history, No. (%) |  |  |  |
| Yes | 313 (4.50) | 272 (4.44) | 41 (5.00) |
| No | 6640 (95.50) | 5861 (95.56) | 779 (95.00) |
| Ovarian cancer history, No. (%) |  |  |  |
| Yes | 235 (3.36) | 209 (3.39) | 26 (3.17) |
| No | 6752 (96.64) | 5958 (96.61) | 794 (96.83) |
| Prostate cancer history, No. (%) |  |  |  |
| Yes | 671 (9.65) | 590 (9.62) | 81 (9.83) |
| No | 6285 (90.35) | 5542 (90.38) | 743 (90.17) |
| Sedentary hours, Mean (SD), h | 13.88 (2.96) | 13.92 (2.97) | 13.60 (2.87) |
| Vitamin D intake, age 30s, Mean (SD), mcg | 0.10 (0.33) | 0.10 (0.32) | 0.14 (0.42) |
| Vitamin D intake, teen age, Mean (SD), mcg | 0.17 (0.47) | 0.17 (0.46) | 0.21 (0.51) |
| Vitamin D intake, childhood, Mean (SD), mcg | 0.43 (0.73) | 0.42 (0.72) | 0.49 (0.78) |
| Height, Mean (SD), cm | 163.01 (9.04) | 163.25 (9.11) | 161.26 (8.34) |
| Weight, Mean (SD), kg | 72.10 (14.83) | 72.41 (14.94) | 69.83 (13.74) |
| Lumbar spine BMD, Mean (SD) | 0.95 (0.18) | 0.96 (0.18) | 0.88 (0.17) |
| Femoral neck BMD, Mean (SD) | 0.72 (0.13) | 0.73 (0.13) | 0.66 (0.11) |
| Total hip BMD, Mean (SD) | 0.88 (0.16) | 0.89 (0.16) | 0.80 (0.14) |
| Ever fracture before baseline, No. (%) |  |  |  |
| Yes | 3463 (44.67) | 2926 (42.81) | 537 (58.50) |
| No | 4290 (55.33) | 3909 (57.19) | 381 (41.50) |
| Number of siblings, Mean (SD) | 4.40 (3.35) | 4.42 (3.37) | 4.24 (3.24) |

<sup>^</sup> Fracture: first incidence of any of the hip, spine, pelvis, shoulder, ribs and forearm fractures over 19 years of follow-up

### Data Pre-Processing and Feature Engineering

We performed several data cleaning and pre-processing steps. First, data quality and missingness were assessed. Missing data was imputed using the bag imputation method. Several feature engineering steps were employed. All continuous features were standardized. We used one-hot encoding for dealing with categorical features, where a binary feature is formed for each level of the categorical feature. For example, one-hot encoding was used for the menopause status as it had three categories: non-menopausal, menopausal, and both ovaries removed. In some cases, the levels of categorical features were combined to form a single binary feature. For example, race had four categories (White, Black, Asian, Other). The category “non-White” was formed by collapsing “Black”, “Asian”, and “Other” while “White” was left unchanged. In addition, features with near-zero variance were excluded from the analysis.

### Machine Learning Algorithms

In this study, we evaluated seven ML algorithms that fall under different categories: standard logistic regression, penalized approach (LASSO, Elastic Net), tree-based approach (Random Forest, Decision Tree), boosted approach (extreme gradient boosting, XGBoost), and deep learning approach (Neural Network).

### Statistical and machine learning Analysis

The descriptive summary were reported as mean (SD) or median (Q1, Q3) for continuous features and as n (%) for categorical features.

The data were randomly split into a training set (n = 5,426; 70%) and a testing set (n = 2,327; 30%). The data split was stratified by fracture status to ensure a similar proportion of fractures in both the training and testing datasets. ML algorithms were then trained on the training dataset, and the performance was evaluated on the testing data. The previously mentioned seven ML algorithms were developed based on the training dataset. The hyperparameters were tuned using a ten-fold nested cross-validation loop with five repetitions. The 20 randomly generated hyperparameters were evaluated in the inner loop, and the model’s performance was assessed in the outer loop in terms of ROC AUC. The Bayesian adaptive approach was used to optimize the hyperparameter tuning process. Once optimal parameter combinations had been selected, the performance of each algorithm was evaluated on the test dataset in terms of ROC_AUC. The ROC_AUC values, along with the 95% confidence intervals (CI), were reported.

The best-performing algorithm was then subsequently used on the whole dataset. Then, a SHAP analysis was performed to quantify the importance of the features. For the SHAP analysis, we collapsed the contribution of three features created through one-hot encoding and named it “Menopause”. Also, we collapsed the contribution of fractures that occurred before the baseline and fractures reported at baseline and named it as “Previous fracture”. We reported the 15 most important features. Thereafter, a SHAP-interaction analysis was performed to investigate how the five most important features interacted with other features. All analyses were conducted using R version 4.3.2^32^.

## Results

In total, 7,753 participants aged 50 years or older were included in the analysis. Most of the participants were female (5,566; 72%), and the mean age was 67 (SD=8.56) years. The features included in the analysis by fracture status were summarized in Table 1.

After pre-processing, 55 (out of 73) features were included to train the ML algorithms. The optimized tuned hyperparameters for all algorithms are given in the Supplementary Table 1. The performance of the ML algorithms, as measured by the ROC_AUC based on the test dataset, is presented in Table 2. The ROC_AUC values for all algorithms were comparable and over 0.65. The XGBoost had the highest ROC_AUC value of 0.700 (95% CI: 0.669, 0.732), while the Decision Tree had the lowest 0.656 (95% CI: 0.624, 0.689). The LASSO and Elastic Net had similar ROC_AUC values of 0.696 (95% CI: 0.664, 0.726) and 0.694 (95% CI: 0.660, 0.726), respectively. The Neural Network and Random Forest also had similar ROC_AUC values of 0.684 (95% CI: 0.649, 0.716) and 0.686 (95% CI: 0.652, 0.719), respectively.

**Table 2:** Performance of the machine learning algorithm on the test dataset.

| ML Algorithm | ROC_AUC (95% CI) |
| --- | --- |
| LASSO | 0.696 (0.664, 0.726) |
| Elastic Net | 0.694 (0.660, 0.726) |
| Neural Net | 0.684 (0.649, 0.716) |
| Random Forest | 0.686 (0.652, 0.719) |
| XGBoost | 0.700 (0.669, 0.732) |
| Logistic Regression | 0.689 (0.654, 0.722) |
| Decision Tree | 0.656 (0.624, 0.689) |

We identified the 15 most important features and their corresponding SHAP values for the prediction of fragility fractures through SHAP analysis (Figure 1). The total hip BMD had the highest SHAP value of 0.119, followed by lumbar spine BMD (0.107), previous fracture (0.102), femoral neck BMD (0.090), age (0.061), menopause status (0.058), weight (0.043), osteoporosis status (0.042), walk status (0.026), vitamin D intake, teen age (0.024), sex (0.024), osteoporosis history (0.023), vitamin D intake/day (0.022), calcium intake/day (0.021) and sedentary hours (0.020) (Figure 1). The SHAP analysis for total hip BMD indicated that the SHAP value increases with low total hip BMD (Figure 1). Similarly, SHAP values increase with low lumbar spine BMD, low femoral neck BMD and vitamin D intake (Figure 1). On the other hand, SHAP value increases with age, weight, previous fracture, osteoporosis, osteoporosis history and menopausal status (Figure 1). SHAP value was higher for females compared to males (Figure 1).

We also investigated how the first five important features, total hip BMD, lumbar spine BMD, previous fracture, femoral neck BMD, and age interacted with other features (Supplementary Tables 2.1-2.5). We found that the total hip BMD interacted the most with femoral neck BMD, with a SHAP value of 0.014 (Supplementary Table 2.1). The interaction plot between total hip and femoral neck BMDs indicated that SHAP value increases with low value of both total hip and femoral neck BMDs (Figure 2A). The other features total hip BMD interacted with were weight, lumbar spine BMD, previous fracture, calcium intake, age, and menopause (Supplementary Table 2.1). Lumbar spine BMD interacted the most with weight (Supplementary Table 2.2) followed by total hip BMD, previous fracture femoral neck BMD, calcium intake and age (Supplementary Table 2.2). The SHAP value increased with low lumbar spine BMD, and either lower or higher weight (Figure 2B). The first five features femoral neck BMD interacted with were total hip BMD, previous fracture, calcium intake/day, lumber spine BMD, and weight (Supplementary Table 2.3). Similarly, the first five features previous fracture interacted with were femoral neck BMD, lumbar spine BMD, vitamin D intake/day, total hip BMD and height. The SHAP value increases with previous fracture and low femoral neck BMD (Figure 2C). Lastly, age interacted the most with lumbar spine BMD (Supplementary Table 2.5). The SHAP value increases with low lumbar spine BMD and either lower or higher age (Figure 2D).

**Figure 1:**
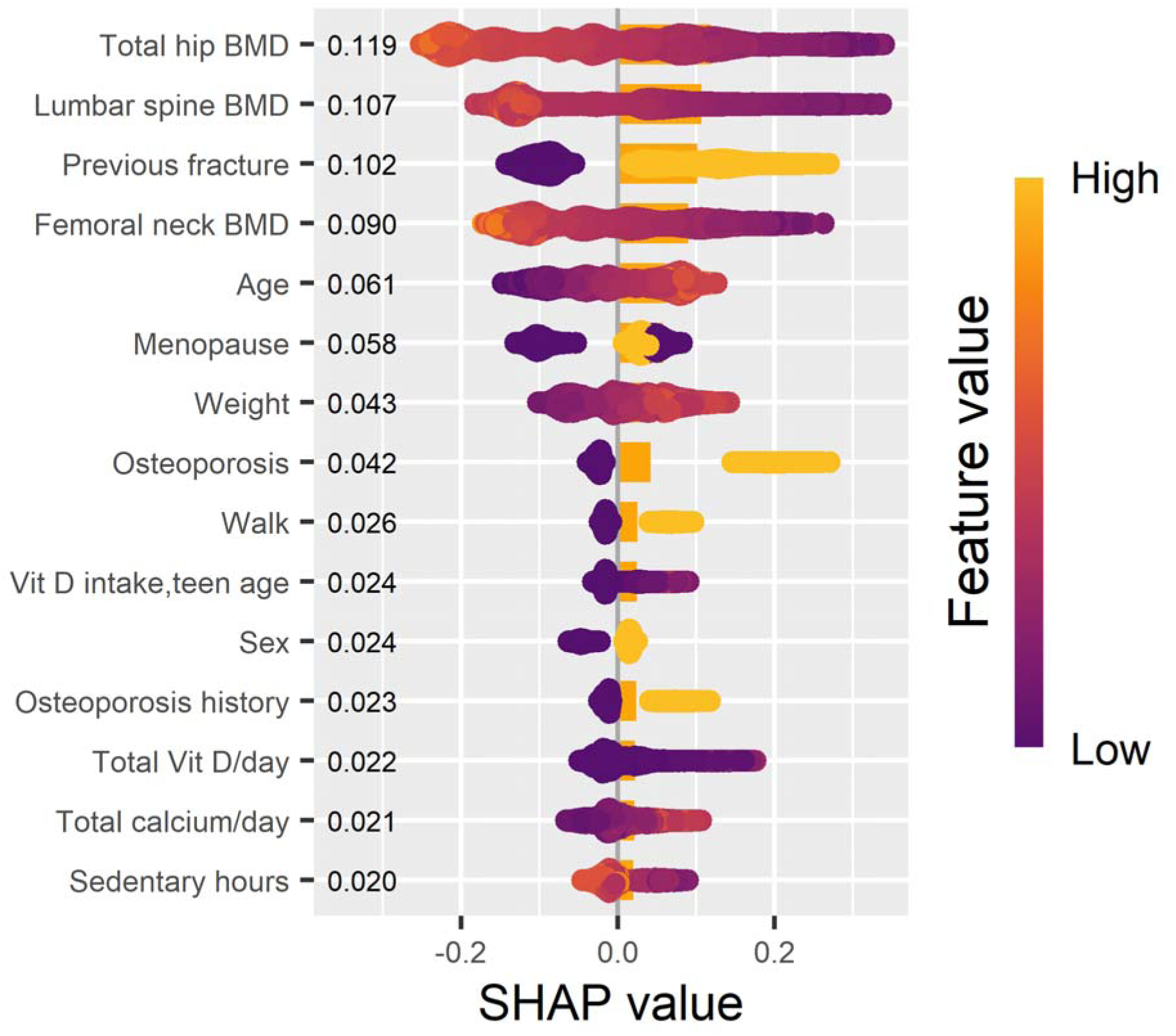
**First 15 important features and the corresponding SHAP values**

**Figure 2:**
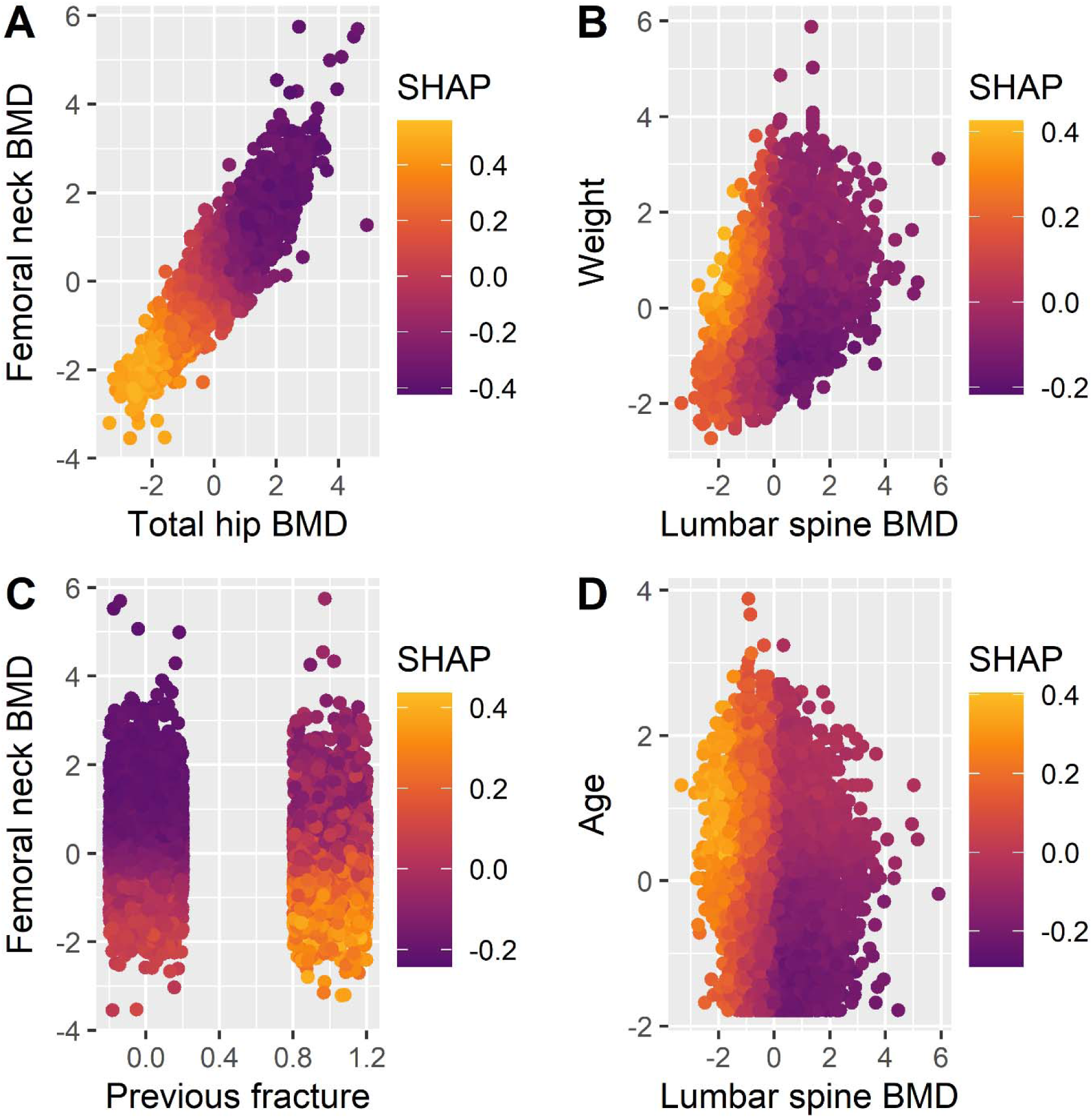
Results of the interaction between the first five important features and the feature with which they had the highest interaction. (One figure for total hip BMD and femoral neck BMD, as these two had the highest interaction)

## Discussion

In this study, we investigated the performance of seven ML algorithms and evaluated them with respect to their ability to predict fragility fractures using the data from the CaMos, where the incidence of fragility fracture (first occurrence of any of hip, spine, pelvis, shoulder, ribs and forearm) was measured over 19 years of follow-up^31^. We found that the XGBoost performed slightly better than all other algorithms in the prediction of fragility fractures.

Fragility fractures can occur at different skeletal sites, including the hip, spine and pelvis ^2^ but we have limited evidence considering the totality of multiple sites. Vries et al. investigated ML algorithms for major osteoporotic fractures, evaluating the incidence of fractures at any of the hip, wrist, spine and humerus, among patients who had a prior fracture^23^. The authors found that Cox regression performed better than Random Forest and deep learning algorithms^23^. This study considered the time to the occurrence of any major osteoporotic fractures^23^, while our study measured the occurrence of fractures as categorical (yes/no).

Researchers have largely focused on ML algorithms for the prediction of hip fracture^20^ due to their severe consequences on the lives of older adults^6,7^. Kruse and colleagues demonstrated that XGBoost was the best method for predicting hip fractures among men^25^. Likewise, Kong et al. reported that the CatBoost, a variation of the XGBoost, was superior compared to logistic regression and support vector machine^26^, while Engels and colleagues^33^ found that the performance of the XGBoost and logistic regression models was almost similar for the prediction of hip fractures. These findings are all in line with the findings from our study.

Moreover, our studied ML algorithms achieved almost similar discriminative performance, measured by ROC_AUC, compared to other studies. For instance, Engels et al., when investigating hip fracture prediction models, demonstrated that both XGBoost and logistic regression had an ROC_AUC value of 0.703 and 0.704, respectively^33^. Similarly, Kong et al found that the CatBoost algorithm yielded an AUC value of 0.688^26^. Furthermore, the ROC_AUC value of the XGBoost algorithm from our study (0.700) was slightly better than the FRAX tool (ROC_AUC = 0.68)^17^ and yielded almost similar performance (AUC = 0.700) as Garvan, another fracture prediction tool^17^.

It is important to note that the superiority of the XGBoost algorithm in this study was marginal, as the next best performing algorithm, LASSO, had an ROC_AUC value of 0.696, a difference of only 0.004. Similar findings were also demonstrated in other studies^26,34^.

We adopted an explainable approach and identified important features and their contribution to the prediction of fragility fractures. The SHAP analysis assesses the contribution of each feature to the prediction while considering all other features. We identified the fifteen most important features, including total hip BMD, lumbar spine BMD, previous fracture, femoral neck BMD, age, menopause, weight, osteoporosis status, osteoporosis history and vitamin D intake. Similarly, from their SHAP analysis, Kong et al. demonstrated that BMDs (total hip, femoral neck and lumbar) were the most important features for the prediction of hip fractures^26^. Age and BMI were also found to be important features in their work, which is similar to this study^26^. Some of the features we identified are known risk factors for fragility fractures, as previous epidemiological research has shown that characteristics such as age, female sex, BMDs, and osteoporosis status are important risk factors for fragility fractures^2,34^.

We also demonstrated how the magnitude of individual features contributes to the prediction of fragility fractures. For example, low values of BMDs (total hip, femoral neck, and lumbar spine) and low intake of vitamin D and calcium increased the risk of fragility fractures. Individuals with osteoporosis or a family history of osteoporosis or previous fractures were at high risk of fracture. Our study further demonstrated that menopausal women were vulnerable to fragility fractures. Calcium and vitamin D intake will be helpful to reduce the risk of fractures, as low intake increases this risk.

We investigated the feature-feature interactions through SHAP analysis for the five most important features. We found that total hip BMD interacted with many factors, including femoral neck BMD, weight, sex, daily calcium intake, menopause status, and age. It had the highest interaction with femoral neck BMD, wherein individuals with low hip BMD and low femoral neck BMD were found to be at a higher risk of fragility fractures. Lumbar spine BMD had the highest interaction with weight and age. Individuals with low lumbar spine BMD and with either higher or lower weight or older age have an elevated risk of fragility fractures. Similarly, individuals with low femoral neck BMD and previous fractures were at greater risk of fragility fractures. These findings are valuable as they will be helpful to identify a particular subset of individuals who are at high risk for fractures, potentially prompting increased screening for this population. Similar to our findings, Kong and colleagues also found that total hip BMD interacts with age to increase the risk of fracture incidence^26^. Kong et al found additional interactions between total hip BMD and factors such as lumbar trabecular bone score and low serum creatinine^26^, which were not considered in this study.

## Strengths and Limitations

This study has several strengths. First, in this study, we used the data from a population-based longitudinal study where fracture incidence was measured over a very long follow-up period^31^. Secondly, we considered a large number and wide range of features from various stages of life to develop the ML algorithms. Finally, we identified important features and their interplay to ascertain how they modulate fragility fracture prediction.

This study also had several limitations. First, the incidence of fractures was mostly self-reported, but subsequently confirmed by medical reports. Most of the participants were female. Finally, we did not include any biomarkers or omics data in this study.

## Conclusion

This study developed and compared the predictive performance of seven ML algorithms to predict the incidence of fragility fractures, utilizing the data from a longitudinal prospective cohort study. We found that the XGBoost algorithm performed slightly better than other tested algorithms. SHAP analysis not only identified key risk factors such as BMD, vitamin D intake, and age that increase the risk of fragility fractures, but also how these important features interact with other features, an area of research seldom explored before.

## Data Availability

Data are not available to share

---

**Supplementary Table 1:** Results of hyperparameter tuning.

| Algorithm | Hyperparameters and their optimal values |
| --- | --- |
| LASSO | penalty =0.00285,mixture = 1 |
| Elastic net | penalty =0.00234,mixture =0.65 |
| XGBoost | mtry = 7, trees = 1572, min_n = 4, tree_depth = 8, learn_rate = 0.00165, loss_reduction = 0.000984, sample_size = 0.168, stop_iter = 4 |
| Decision tree | tree_depth= 6, cost_complexity = 0.00000183, min_n = 22 |
| Random forest | mtry = 2, min_n = 37, trees = 762 |
| Neural network | hidden_units = 7, penalty =0.0650, epochs =10 |
| Logistic regression | NA |

**Supplementary Table 2.1:** Results of SHAP interaction analysis between total hip BMD and other features with SHAP values>=0.002.

| Feature | Total Hip BMD |
| --- | --- |
| Femoral neck BMD | 0.014 |
| Weight | 0.013 |
| Lumbar spine BMD | 0.008 |
| Previous fracture | 0.006 |
| Total calcium/day | 0.005 |
| Age | 0.004 |
| Menopause | 0.004 |
| Height | 0.004 |
| Osteoporosis history | 0.003 |
| Lives alone | 0.002 |
| Drinks/year | 0.002 |
| Total Vit D/day | 0.002 |
| Vit D intake, teen age | 0.002 |
| Sex | 0.002 |
| Sedentary hours | 0.002 |
| No. of siblings | 0.002 |
| Education | 0.002 |
| Cancer history | 0.002 |
| Thyroid | 0.002 |

**Supplementary Table 2.2:** Results of SHAP interaction analysis between lumbar spine BMD and other features with SHAP values>=0.002.

| **Feature** | **Lumbar spine BMD** |
| --- | --- |
| Weight | 0.010 |
| Total hip BMD | 0.008 |
| Previous fracture | 0.008 |
| Femoral neck BMD | 0.008 |
| Total calcium/day | 0.007 |
| Age | 0.006 |
| Osteoporosis history | 0.005 |
| Menopause | 0.004 |
| Osteoporosis | 0.004 |
| Drinks/year | 0.004 |
| Height | 0.003 |
| Total Vit D/day | 0.003 |
| Vit D intake, teenage | 0.002 |
| No. of siblings | 0.002 |
| Smoking | 0.002 |
| Sedentary hours | 0.002 |
| Education | 0.002 |
| Sleep | 0.002 |
| Vit D intake, childhood | 0.002 |

**Supplementary Table 2.3:** Results of SHAP interaction analysis between femoral neck BMD and other features with SHAP values>=0.002.

| **Feature** | **Femoral neck BMD** |
| --- | --- |
| Total hip BMD | 0.014 |
| Previous fracture | 0.009 |
| Total calcium/day | 0.008 |
| Lumbar spine BMD | 0.008 |
| Weight | 0.007 |
| Menopause | 0.005 |
| Live alone | 0.004 |
| Age | 0.004 |
| Height | 0.004 |
| Sedentary hours | 0.003 |
| Education | 0.003 |
| Vit D intake, childhood | 0.002 |
| Total Vit D/day | 0.002 |
| Vit D intake, teenage | 0.002 |
| Osteoporosis history | 0.002 |
| Sex | 0.002 |
| Drinks/year | 0.002 |
| No. of siblings | 0.002 |
| Sleep | 0.002 |

**Supplementary Table 2.4:** Results of SHAP interaction analysis between previous fracture and other features with SHAP values>=0.002.

| Feature | Previous fracture |
| --- | --- |
| Femoral neck BMD | 0.009 |
| Lumbar spine BMD | 0.008 |
| Total Vit D/day | 0.006 |
| Total hip BMD | 0.006 |
| Height | 0.005 |
| Age | 0.004 |
| Total calcium/day | 0.004 |
| Walk | 0.004 |
| Menopause | 0.004 |
| Education | 0.003 |
| Drinks/year | 0.003 |
| No. of siblings | 0.003 |
| Weight | 0.002 |
| Vit D intake, childhood | 0.002 |
| Sex | 0.002 |
| Sedentary hours | 0.002 |
| Vit D intake, age 30s | 0.002 |
| Osteoporosis | 0.002 |
| Vit D intake, teenage | 0.002 |
| CVD history | 0.002 |

**Supplementary Table 2.5:** Results of SHAP interaction analysis between age and other features with SHAP values>=0.002.

| Feature | Age |
| --- | --- |
| Lumbar spine BMD | 0.006 |
| Previous fracture | 0.004 |
| Total hip BMD | 0.004 |
| Total calcium/day | 0.004 |
| Femoral neck BMD | 0.004 |
| Weight | 0.003 |
| No. of siblings | 0.003 |
| Menopause | 0.003 |
| Moderate Activity | 0.002 |
| Sex | 0.002 |
| Total Vit D/day | 0.002 |

## Notes

### Competing Interest Statement

Acharya and Drs. Borhan, Thabane, Hanely, Berger, and Morin declared no conflict of interest. Dr. Papaioannou reported receiving honoria from Amgen and funding from Osteoporosis Canada. Dr. Goltzman reported receiving funding from the Canadian Institute of Health Research (CIHR), one-time royalties from UpToDate, one consulting fee from Biosyent, patents: 2457928(Canada), 60/384122(USA); 2343713(Canada) issued to the McGill University, and provided clinical expert assessment of Burosumab for treatment of X-linked Hypophosphatemia(XLH). Dr. Adachi reported receiving funding from CIHR, Eli Lily, Merck, Procter & Gamble, Sanofi, Amgen, consulting fees and honoraria from Amgen. Dr. Raina reported receiving funding from the Canadian Institute of Health Research (CIHR) and the Canada Foundation for Innovation and being involved with the WHO working group on life course.

### Funding Statement

Dr. Borhan received partial funding through the OC-CaMos fellowship from Osteoporosis Canada to conduct this study.

### Author Declarations

Hamilton Integrated Research Ethics Board (HiREB) gave ethical approval of this work.

### Summary of Updates

The manuscript has been formatted for another journal.

